# A Large-Scale, International Cross-Sectional Survey of Published Pediatrics Authors: Perceptions of Complementary, Alternative, and Integrative Medicine

**DOI:** 10.1101/2024.10.08.24315106

**Authors:** Jeremy Y. Ng, Gursimran Deol, Dennis Anheyer, Holger Cramer

**Affiliations:** Institute of General Practice and Interprofessional Care, University Hospital Tübingen, Tübingen, Germany; Bosch Health Campus, Stuttgart, Germany; Department of Psychology and Psychotherapy; Chair of Methodology and Statistics in Psychology, Universität Witten/Herdecke, Witten Germany

**Keywords:** complementary and alternative medicine, integrative medicine, pediatric, pediatrics, perceptions

## Abstract

**Background:** The use of complementary, alternative, and integrative medicine (CAIM) is commonly used among pediatric patients for various conditions. Pediatrics clinicians and researchers’ perceptions towards the incorporation of CAIM therapies have varied. This study aims to investigate the perceptions of both pediatrics researchers and clinicians regarding CAIM.

**Methods:** We conducted a large-scale, international cross-sectional online survey with published pediatrics authors who have published their work in pediatrics medical journals that are indexed in MEDLINE. In total, 34 494 researchers and clinicians were invited to complete the survey. The survey recorded respondents’ perceptions on various CAIM therapies. Descriptive statistics were generated from the quantitative survey results. A thematic analysis was conducted for responses to open ended questions.

**Results:** In total, 731 pediatrics clinicians and/or researchers responded to the survey, with about half of the respondents being faculty members/principal investigators (56.10%) and/or clinicians (43.45%) and from the Americas (46.56%) or Europe (30.53%). Over half of the respondents viewed mind-body therapies favourably (62.01%) and the fewest respondents held favourable perceptions of biofield therapies (6.98%). Respondents agreed or strongly agreed that there is value in conducting further research on CAIM therapies (85.52%) and disagreed or strongly disagreed that they felt comfortable recommending most CAIM therapies to patients (64.83%). A thematic analysis of our findings demonstrates that many pediatrics clinicians and/or researchers support further research on CAIM.

**Conclusion:** The findings from this study demonstrate that pediatrics clinicians and researchers have varying perceptions towards CAIM therapies. Respondents had the most positive perceptions of mind-body therapies and felt they did not have adequate training on CAIM. Further research is needed to establish more evidence-based educational resources on CAIM.

## Introduction

“Complementary medicine” involves non-mainstream therapies which are used in conjunction with conventional medicine. Conversely, “alternative medicine” refers to non-mainstream therapies being used instead of conventional medicine. “Integrative health” is the coordinated use of both conventional and complementary approaches [1,2]. For the purpose of this study, we will refer to these terms collectively as complementary, alternative, and integrative medicine (CAIM). Together, CAIM refers to a diverse range of health systems, modalities, practices, and products that fall outside of the domain of conventional medicine but can be used in place of or in conjunction with conventional medical treatment to address health conditions [3–6]. The National Health Interview Survey in 2012 found that CAIM therapies are used by more than one in every ten pediatric patients [7]. The prevalence increases to more than one in every two patients among children with chronic diseases [7–10]. CAIM therapies are also frequently used by children and adolescents with chronic pain, infections, cancers, dermatological conditions, gastrointestinal conditions, hematological conditions neurological conditions, and respiratory conditions [11,12]. CAIM modalities common among pediatric patients include acupuncture, herbal medicine, homeopathy, massage, naturopathy, therapeutic touch, relaxation techniques and imagery [13,14]. As younger pediatric patients are unable to provide informed consent, parents or guardians are often the primary decision makers for the pediatric population. Positive parental perceptions of CAIM and sociocultural beliefs likely play a role in how parents or guardians acquire information about CAIM and elect to use it [15–17].

Attitudes and perceptions towards CAIM within the pediatrics medical community are varied. Pediatricians generally indicate willingness to incorporate CAIM modalities into treatment plans and acknowledge the importance of knowing which CAIM therapies are utilized by their patients [18–20]. Despite this, a major proportion of clinicians do not ask questions regarding their patients’ use of CAIM [19,21]. Furthermore, most pediatrics clinicians express a lack of knowledge on the topic and a need for further education [19–22]. Pediatrics clinicians also remain concerned about the lack of evidence supporting the safety and efficacy of some CAIM modalities [19,20,22]. This divide in opinion creates barriers for clinicians and researchers who want to explore and investigate the potential benefits of CAIM practices [23]. It also contributes to inconsistencies in approaches aimed at integrating CAIM into conventional pediatrics healthcare [24].

Given the increasing interest in CAIM and polarized opinions on their therapeutic potential within the pediatrics medical community, it is important to obtain a comprehensive understanding of the perceptions of pediatrics clinicians and researchers on CAIM modalities and practices. As such, this survey aims to investigate the perceptions of both pediatrics researchers and clinicians regarding CAIM.

## Methods

### Transparency Statement

Clearance was obtained from the University Hospital Tübingen Research Ethics Board before commencing this research project (REB Number: 389/2023BO2). The study protocol was registered and made accessible via a platform called the Open Science Framework (OSF) before participant recruitment began [25]. OSF was also used to share study materials and raw data, which can be found at: https://doi.org/10.17605/OSF.IO/Z6U4X.

### Study Design

This study was an anonymous, online, cross-sectional survey of published pediatrics authors who have published in medical journals indexed in MEDLINE [26].

### Sampling Framework

A sample of corresponding authors from all articles published between December 1, 2018 and July 1, 2023 were selected from a sample of pediatrics journals as found here: https://journal-reports.nlm.nih.gov/broad-subjects/. National Library of Medicine (NLM) IDs associated with the selected journals were extracted [27]. A search strategy was generated using these NLM IDs and subsequently, run on OVID Medline. The list of PMIDs compiled from articles identified in the search was exported as a .csv file and inputted into an R script to extract the authors’ names and email addresses. The R script has been designed in accordance with the easyPubMed package [28]. Authors who had published any type of manuscript were included.

### Participant Recruitment

Published pediatrics authors were identified using the sampling framework to participate in this study by completing the survey. Duplicate email addresses were removed. The survey was closed, meaning that only invitees had access to this survey, and they were instructed not to share the survey link with anyone else. Emails were sent out to the prospective participants via SurveyMonkey [29]. Numbers of invalid and non-functional emails which bounced back were recorded by SurveyMonkey. The authors in our sample received an initial email on either October 30, 2023 or October 31, 2023. This initial email included a description of the study, the objectives, and a link to access the survey. Clicking on the survey link led the participants to an informed consent form. After the participants provided consent, participants were taken to the survey questions. The survey contained 33 questions in a one-by-one order, spanning over 12 pages. Reminder emails were sent out after the first, second, and third weeks following the initial invitation. The survey was closed on December 19, 2023, exactly 4 weeks after the last reminder was sent out. Financial compensation was not provided for participation. Moreover, there was no requirement for those who were emailed a survey link to partake in this study. Participants who did not wish to answer any given question were able to skip it and no personal identifying information was collected.

### Survey Design

The survey began with a screening question. Respondents were asked several questions covering demographic information and their perceptions of CAIM. The survey questions primarily followed a multiple-choice format and an open-response question. Respondents were able to use the back button to change answers prior to submitting the survey. The survey was pilot tested prior to distribution by two independent CAIM researchers and their feedback was integrated into the survey. Since these researchers are not affiliated with this project and are highly experienced with CAIM, their feedback serves to improve the face validity of this survey. A copy of the survey can be found on the following link: https://osf.io/4d23w.

### Data Management and Analysis

Descriptive statistics were generated from the analysis of the quantitative data and reported. Qualitative data was analyzed using thematic content analysis. A data driven approach was taken to analyze the responses to the open-ended question, “do you have anything else to share about your perceptions of CAIM?”. Open-ended responses were interpreted and assigned a code. These codes were representations of respondent responses and then were further grouped into themes. Themes were created by the authors based on the patterns and commonalities observed in the data. An inductive coding method was used with no specific, underpinning theories to guide analysis. The Checklist for Reporting Results of Internet E-Surveys (CHERRIES) was used to inform the reporting of this survey study [30].

## Results

### Demographics

A total of 39 434 emails were sent out of which 22 412 remained unopened, 12082 were opened and 4394 bounced. The response rate was 1.85% including bounced emails, 2.12% excluding bounced emails, and 6.05% excluding bounced and unopened emails. There were 731 responses which were broken down into 604 fully completed surveys and 127 partially completed surveys, with a completion rate of 82.6%. Respondents took an average of 9 minutes and 37 seconds to complete the survey. Approximately 55.64% (n=365) of the respondents were female and the most common age of respondents was between 35 to 44 (n=180, 27.44%) or 45 to 54 (n=178, 27.13%). Most respondents identified as both a clinician and a researcher (n=391, 55.23%). From the respondents who specified their roles, respondents were primarily faculty members/principal investigators (n=368, 56.10%), clinicians (n=285, 43.45%) and/or scientists in academia (n=162, 24.70%). Most respondents did not identify as being a part of a visible minority (n=545, 82.95%) and were most commonly from the following World Health Organization regions: Americas (n=305, 46.56%) and Europe (n=200, 30.53%). In terms of the respondent’s stage of career and type of research conducted, most respondents had been working for more than 10 years (n=406, 61.89%) and conducted clinical research (n=439, 76.61%). **Table 1** contains a detailed breakdown of respondent demographics. Raw survey data is available at: https://osf.io/43xz5. Crosstabs for key demographic variables can also be found on OSF: https://osf.io/z6u4x/.

**Table 1:**
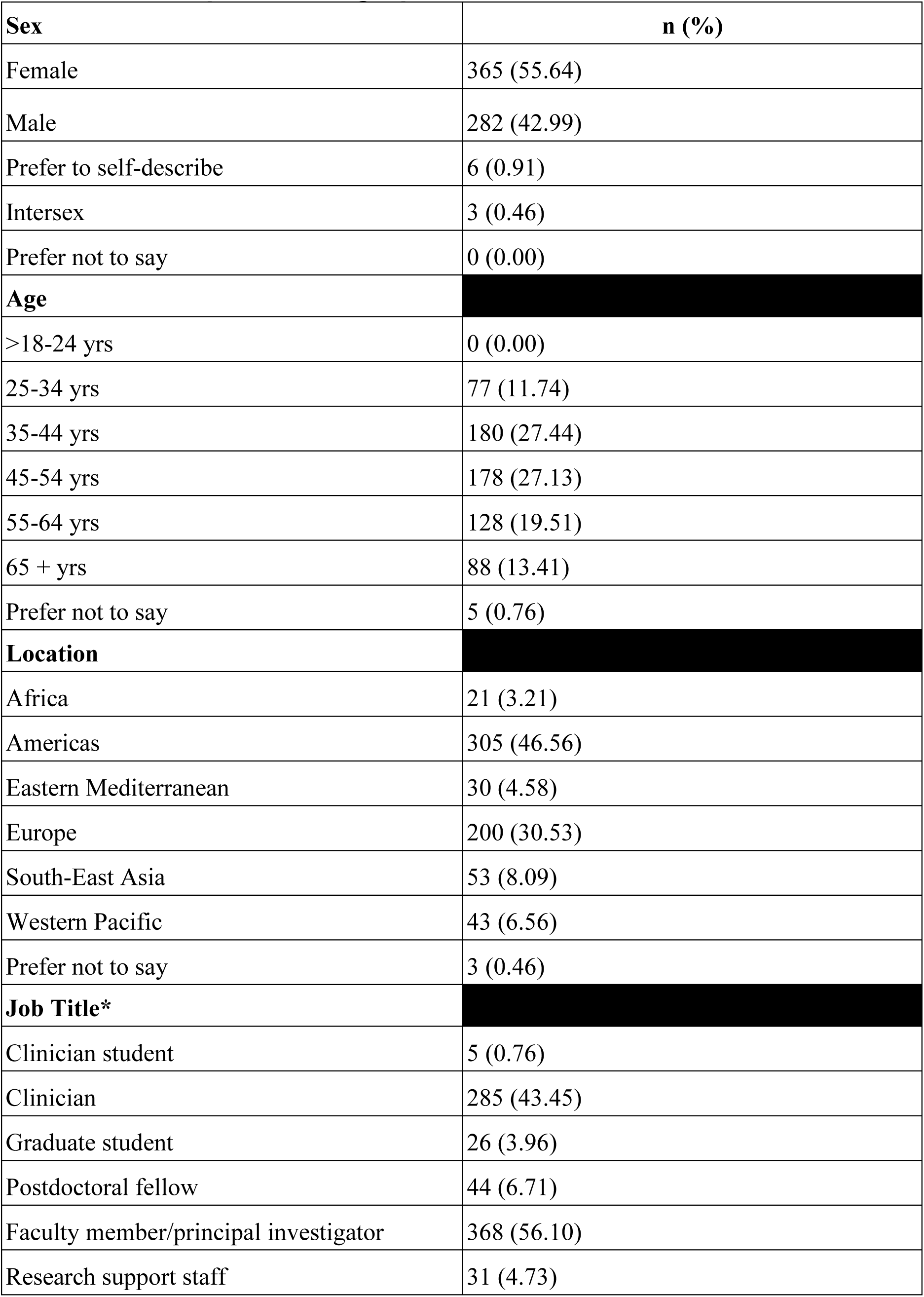
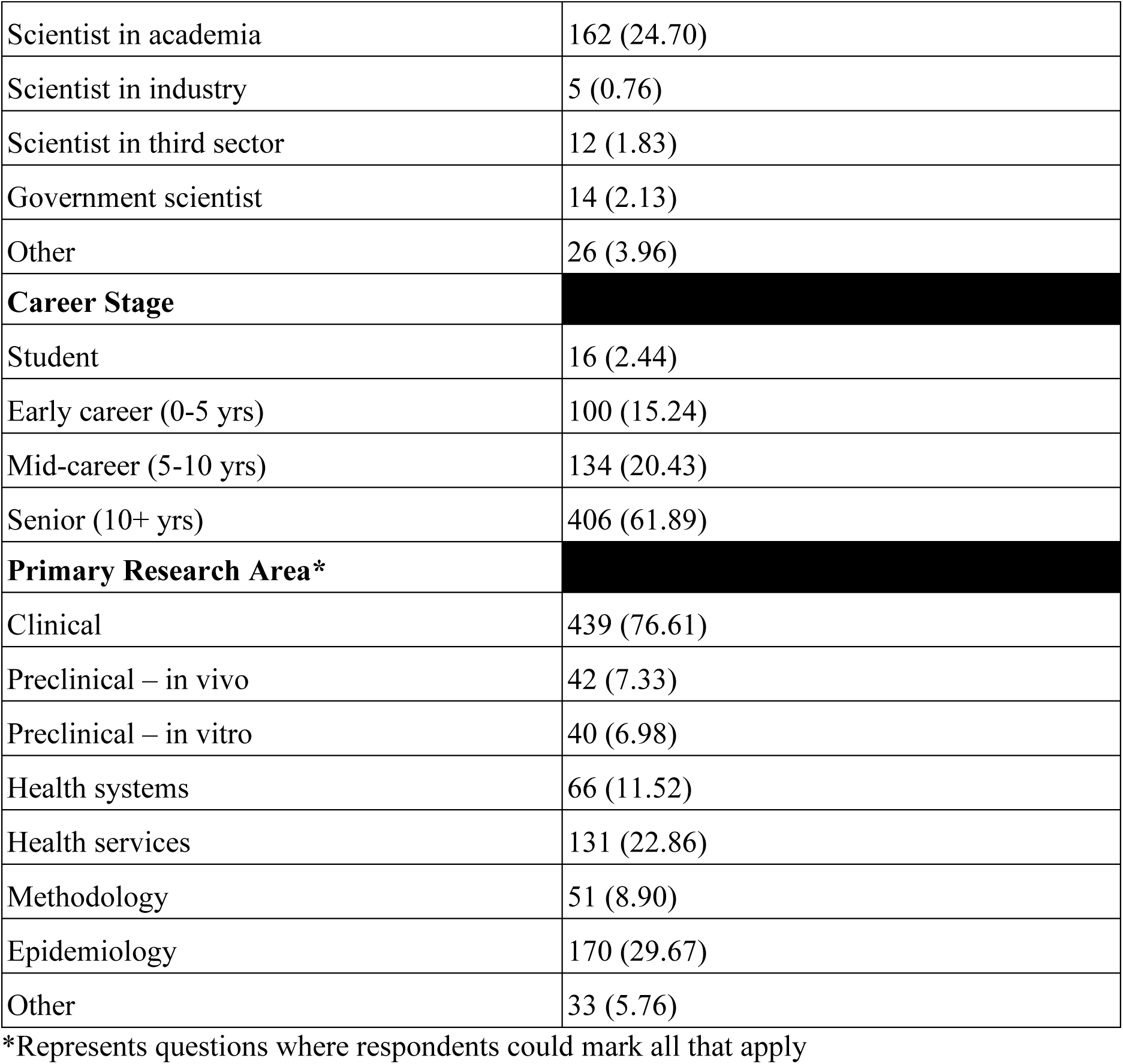
Participant Demographics.

| **Sex** | **n (%)** |
| --- | --- |
| Female | 365 (55.64) |
| Male | 282 (42.99) |
| Prefer to self-describe | 6 (0.91) |
| Intersex | 3 (0.46) |
| Prefer not to say | 0 (0.00) |
| **Age** |  |
| >18-24 yrs | 0 (0.00) |
| 25-34 yrs | 77 (11.74) |
| 35-44 yrs | 180 (27.44) |
| 45-54 yrs | 178 (27.13) |
| 55-64 yrs | 128 (19.51) |
| 65 + yrs | 88 (13.41) |
| Prefer not to say | 5 (0.76) |
| **Location** |  |
| Africa | 21 (3.21) |
| Americas | 305 (46.56) |
| Eastern Mediterranean | 30 (4.58) |
| Europe | 200 (30.53) |
| South-East Asia | 53 (8.09) |
| Western Pacific | 43 (6.56) |
| Prefer not to say | 3 (0.46) |
| **Job Title\*** |  |
| Clinician student | 5 (0.76) |
| Clinician | 285 (43.45) |
| Graduate student | 26 (3.96) |
| Postdoctoral fellow | 44 (6.71) |
| Faculty member/principal investigator | 368 (56.10) |
| Research support staff | 31 (4.73) |
| Scientist in academia | 162 (24.70) |
| Scientist in industry | 5 (0.76) |
| Scientist in third sector | 12 (1.83) |
| Government scientist | 14 (2.13) |
| Other | 26 (3.96) |
| **Career Stage** |  |
| Student | 16 (2.44) |
| Early career (0-5 yrs) | 100 (15.24) |
| Mid-career (5-10 yrs) | 134 (20.43) |
| Senior (10+ yrs) | 406 (61.89) |
| **Primary Research Area\*** |  |
| Clinical | 439 (76.61) |
| Preclinical – in vivo | 42 (7.33) |
| Preclinical – in vitro | 40 (6.98) |
| Health systems | 66 (11.52) |
| Health services | 131 (22.86) |
| Methodology | 51 (8.90) |
| Epidemiology | 170 (29.67) |
| Other | 33 (5.76) |

\*Represents questions where respondents could mark all that apply

### Complementary Alternative and Integrative Medicine

To learn more about CAIM, clinicians and researchers stated they would turn to academic literature (n=544, 88.74%), conferences (n= 263, 42.90%) and/or colleagues (n=215, 35.07%). See **Figure 1** for breakdown of where pediatrics clinicians and researchers would source CAIM information. In a clinical setting, respondents have practiced or recommended the following therapies (in descending order): mind-body (n=181, 45.14%), none (n=148, 36.91%) and biologically based (n=135, 33.67%). Most respondents had received no formal (medical school, residency, etc.) or supplemental training (webinar, conference, etc.) on CAIM therapies. See **Figure 2** for a breakdown of the types of CAIM therapies in which respondents had formal or supplemental training. The percentage of respondents that believed one particular CAIM modality was the most promising for pediatric populations varied: mind-body (n=382, 62.01%), biologically based (n=275, 44.64%), manipulative and body-based (n=145, 23.54%). Patients that were treated by respondents sought counselling for and disclosed the use of biologically based (n=300, 74.26%), mind-body (n= 210, 51.98%) and manipulative and body-based therapies (n= 204, 50.50%). Approximately 55% of respondents (n=220) said that 0-10% of their patient population sought counselling for or disclosed use of CAIM with the majority of respondents having been asked about CAIM occasionally (n=370, 59.97%) as opposed to often (n=97, 15.72%) and never (n=150, 24.31%). **Table 2** contains a detailed breakdown of CAIM research and clinical experiences. Respondents see several benefits and harms to using CAIM. Respondents perceived a lack of scientific evidence for safety and efficacy (n=519, 93.01%) and a lack of standardization in product quality and dosing (n=482, 86.38%) as the greatest challenges with CAIM. Perceived benefits of CAIM included a holistic approach to health and wellness (n=352, 64.83%), expanded treatment options for patients (n=344, 63.35%) and a focus on prevention and lifestyle changes (n=344, 63.35%). **Table 3** contains a detailed breakdown of respondent perceptions as they relate to the challenges and benefits of CAIM.

**Figure 1:**
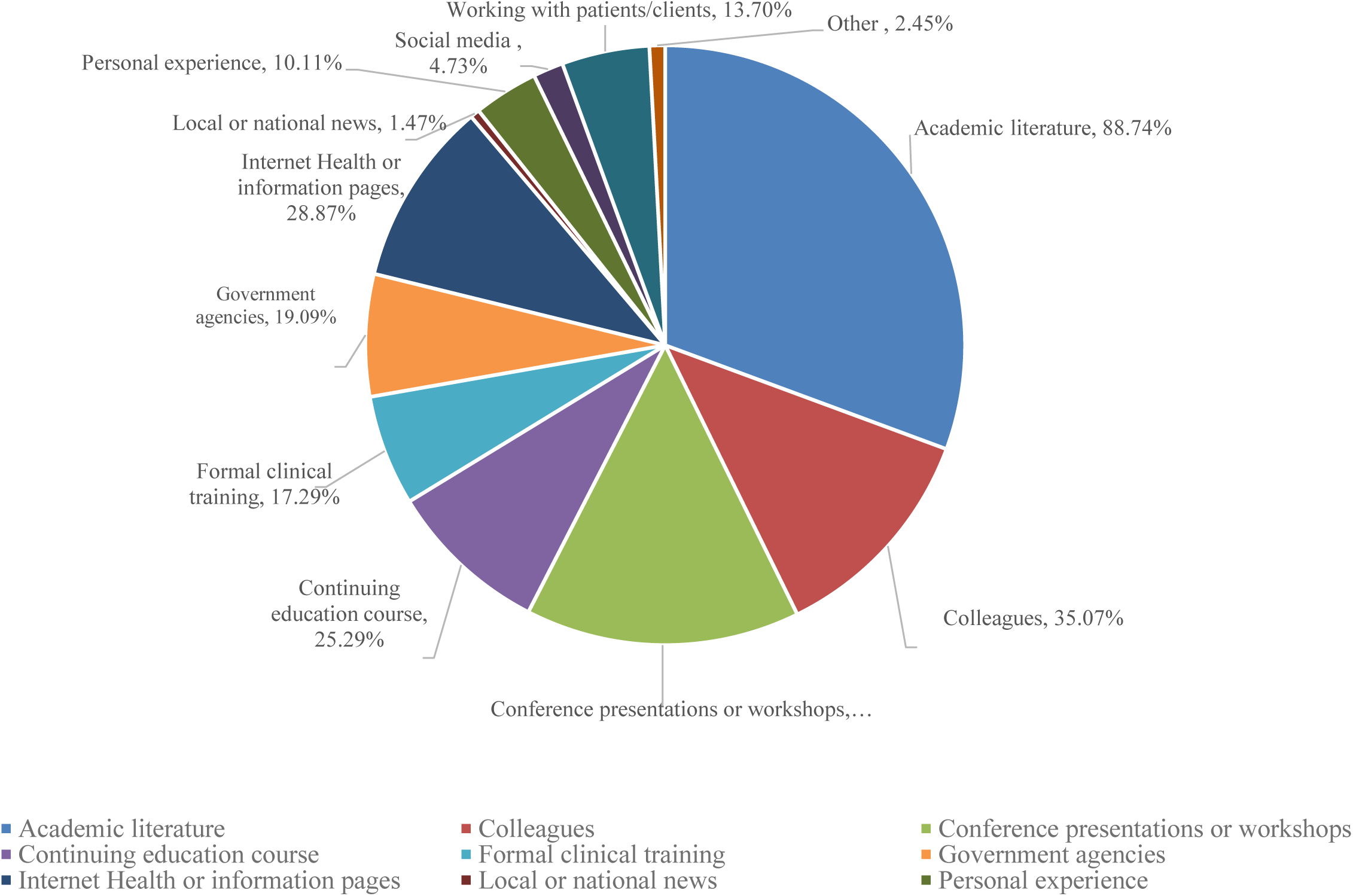
Sources of Information for Pediatrics Clinicians and Researchers

**Figure 2:**
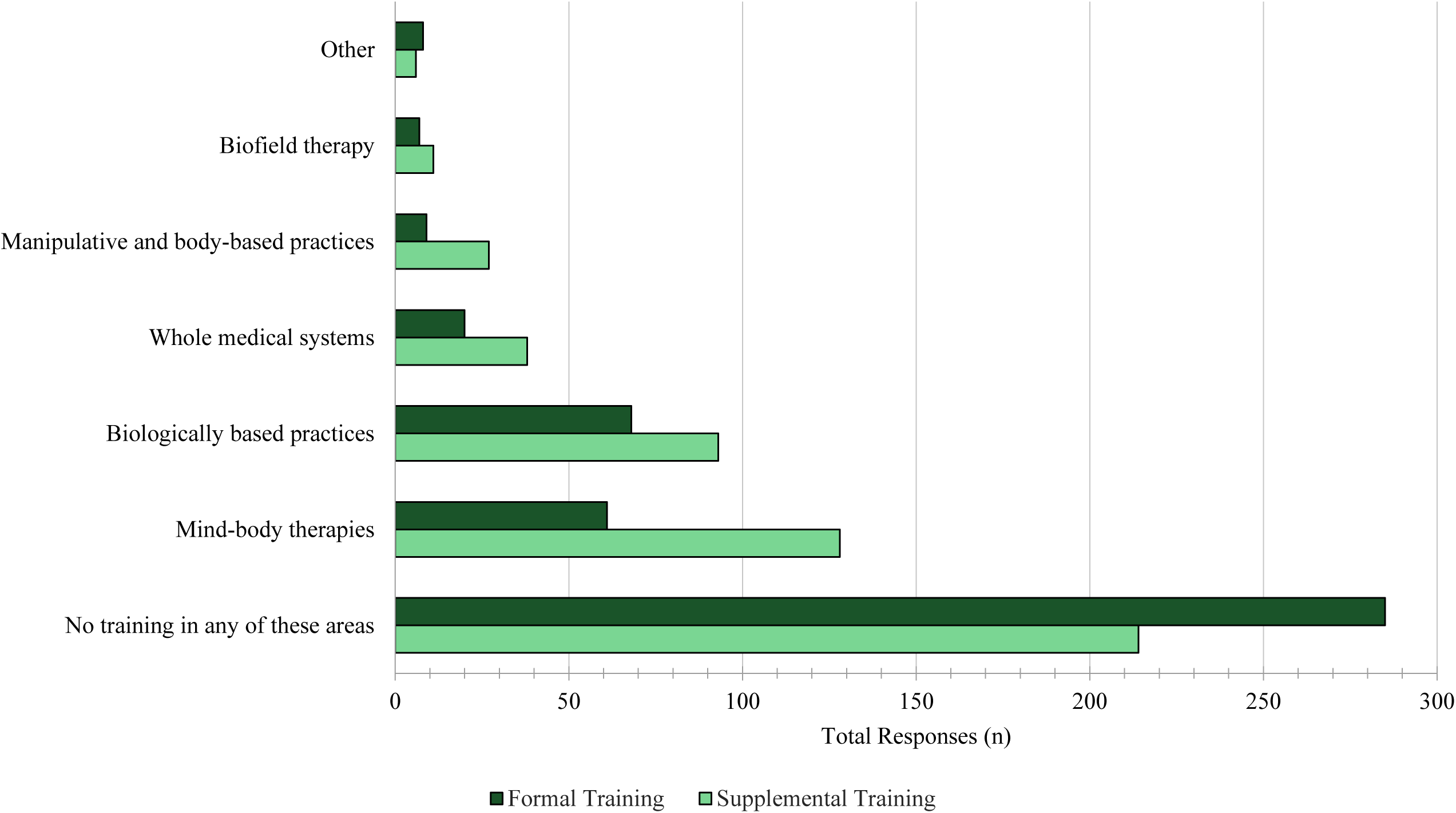
Formal and Supplementary CAIM training in Pediatrics Clinicians and Researchers

**Table 2:** Involvement of CAIM in Clinical Research and Practices.

| **CAIM Research Area** | **n (%)** |
| --- | --- |
| Mind-body therapies | 69 (12.08) |
| Biologically based practices | 101 (17.69) |
| Manipulative and body-based practices | 18 (3.15) |
| Biofield therapies | 9 (1.58) |
| Whole medical systems | 24 (4.20) |
| Never conducted CAIM research | 397 (69.53) |
| Other | 24 (4.20) |
| **Perceived Promising CAIM Therapies** |  |
| Mind-body therapies | 382 (62.01) |
| Biologically based practices | 275 (44.64) |
| Manipulative and body-based practices | 145 (23.54) |
| Biofield therapies | 43 (6.98) |
| Whole medical systems | 131 (21.27) |
| Do not perceive any CAIM categories to be promising | 94 (15.26) |
| Other | 34 (5.52) |
| **Percentage of Patients Counselling and Disclosed Use of CAIM** |  |
| 0-10% | 220 (55.00) |
| 11-20% | 74 (18.50) |
| 21-30% | 50 (12.50) |
| 31-40% | 24 (6.00) |
| 41-50% | 13 (3.25) |
| 51-60% | 9 (2.25) |
| 61-70% | 4 (1.00) |
| 71-80% | 2 (0.50) |
| 81-90% | 3 (0.75) |
| 91-100% | 1 (0.25) |
| **CAIM Areas Patients Counselling and Disclosed Use** |  |
| Mind-body therapies | 210 (51.98) |
| Biologically based practices | 300 (74.26) |
| Manipulative and body-based practices | 204 (50.50) |
| Biofield therapy | 61 (15.10) |
| Whole medical systems | 183 (45.30) |
| I have never had a patient seek counselling or disclose using CAIM | 63 (15.59) |
| **CAIM Practices and Recommendations to Patients** |  |
| Mind-body therapies | 181 (45.14) |
| Biologically based practices | 135 (33.67) |
| Manipulative and body-based practices | 74 (18.45) |
| Biofield therapy | 19 (4.74) |
| Whole medical systems | 46 (11.47) |
| Never practiced or recommended CAIM to patients | 148 (36.91) |
| Other | 8 (2.00) |

**Table 3:** Perceived Challenges and Barriers with CAIM.

| **Perceived Challenges with CAIM** | **n (%)** |
| --- | --- |
| Lack of scientific evidence for safety and efficacy | 519 (93.01) |
| Lack of standardization in product quality and dosing | 482 (86.38) |
| Difficulty in distinguishing legitimate practices from scams or fraudulent claims | 440 (78.85) |
| Limited regulation and oversight | 437 (78.32) |
| Potential interactions with prescription medications | 298 (53.41) |
| Limited integration with mainstream healthcare systems | 299 (53.58) |
| Limited patient education and understanding | 265 (47.49) |
| High cost and lack of insurance coverage | 250 (44.80) |
| Stigmatization and skepticism from healthcare providers and the public | 246 (44.09) |
| Limited availability in certain geographic areas or for certain populations | 204 (36.56) |
| Other | 31 (5.56) |
| **Perceived Benefits with CAIM** | **n (%)** |
| Holistic approach to health and wellness | 352 (64.83) |
| Expanded treatment options for patients | 344 (63.35) |
| Focus on prevention and lifestyle changes | 344 (63.35) |
| Cultural and spiritual relevance for certain populations | 302 (55.62) |
| Empowerment of patients to take control of their own health | 307 (56.54) |
| Potential to address chronic health conditions that conventional medicine has been unable to treat effectively | 270 (49.72) |
| Increased patient satisfaction and well-being | 261 (48.07) |
| Integration with conventional medicine to provide CAIM options | 229 (42.17) |
| Potential cost savings for patients and healthcare systems | 163 (30.02) |
| Fewer side effects than conventional medicine | 152 (27.99) |
| Other | 35 (6.45) |

Participants were asked about their opinions on various prompts and were requested to indicate their level of agreement on a scale ranging from strongly disagree to strongly agree. With regards to CAIM therapies in general, approximately one in three respondents disagreed in feeling comfortable recommending (n=123, 33.79%) and counselling (n=124, 34.07%) patients. Approximately one in three respondents expressed neutrality (neither agreed or disagreed) when asked whether CAIM therapies should be covered by insurance companies (n=178, 31.50%), whether CAIM therapies should be integrated into mainstream medical practice (n=188, 33.39%), if CAIM therapies are safe (n=214, 37.88%) and if CAIM therapies are effective (n=238, 42.20%). Respondents most commonly agreed that clinicians should receive training on CAIM therapies through supplementary (n=292, 51.87%) and formal (n=250, 44.17%) education, more research funding should be allocated to study CAIM therapies (n=218, 38.58%) and there is value in conducting research on CAIM therapies (n=284, 50.18%). Respondents most commonly strongly disagreed with feeling comfortable recommending most CAIM therapies to patients (n=113, 31.04%) and most commonly strongly agreed with ensuring that there is value in conducting research on CAIM therapies (n=200, 35.34%). See **Figure 3** for a detailed breakdown of pediatrics clinicians and/or researchers’ perceptions of CAIM. Please see **Figures 4-8** for detailed breakdown of results by specific CAIM modalities.

**Figure 3:**
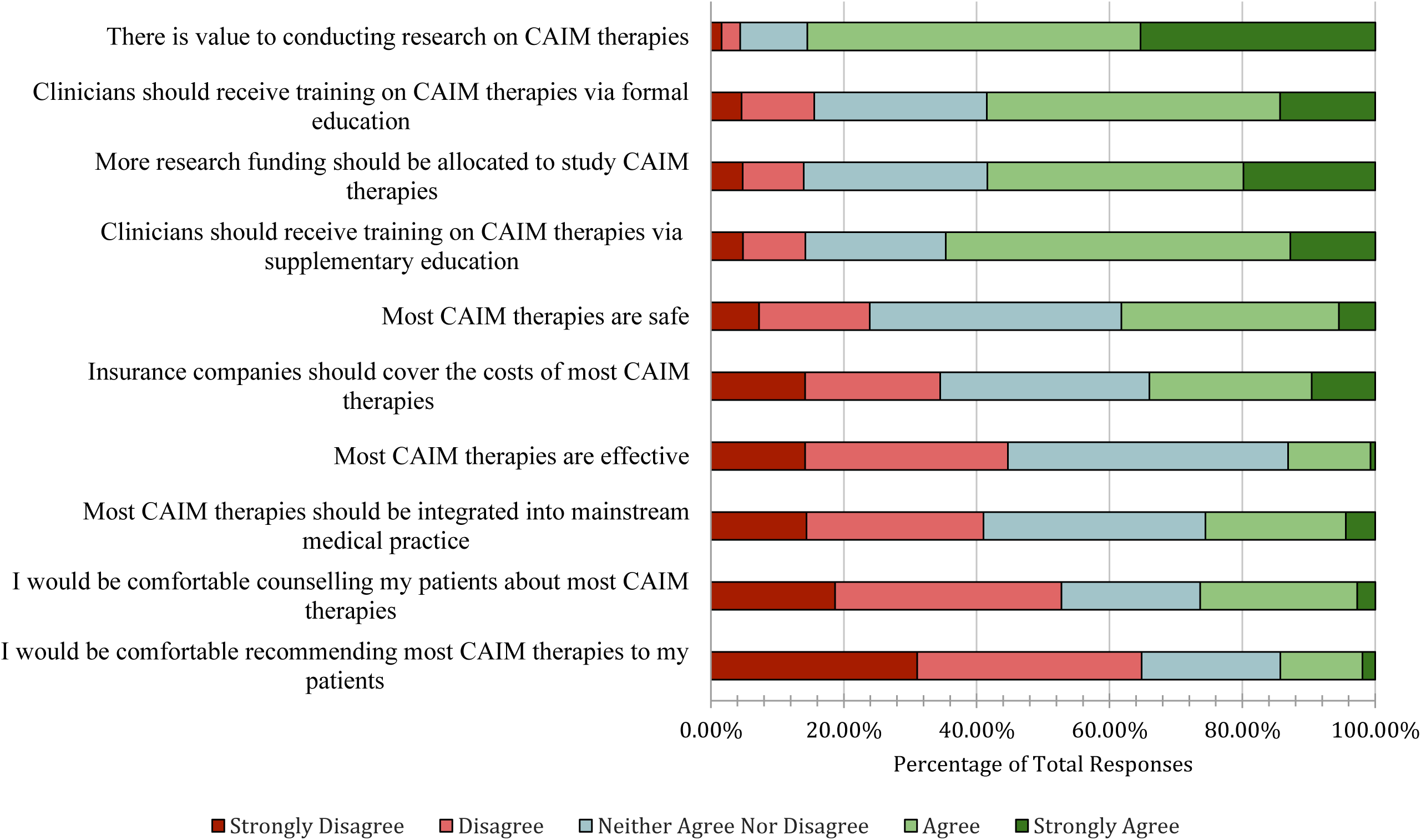
Agreement with the Following Statements Regarding CAIM in General

**Figure 4:**
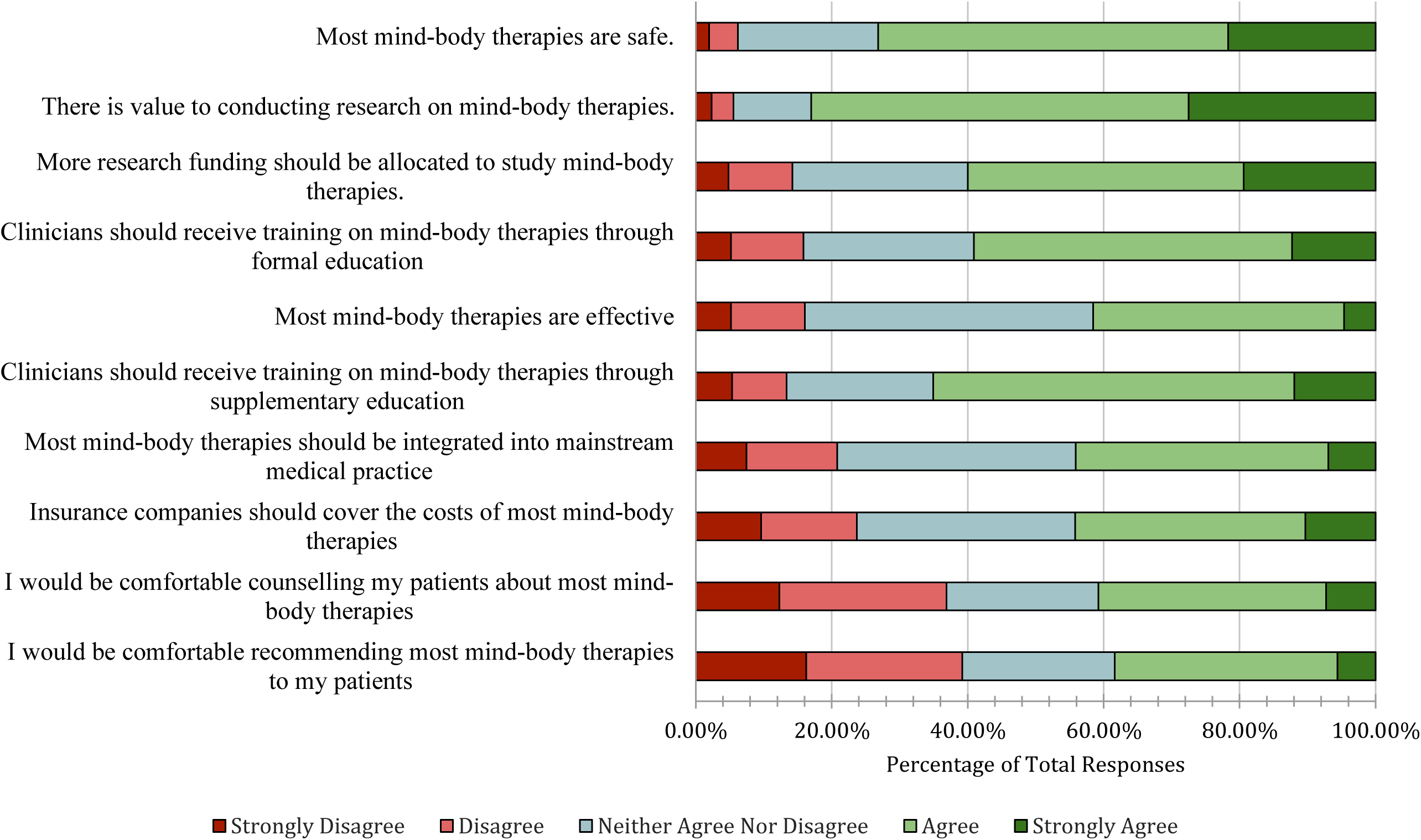
Agreement with the Following Statements Regarding Mind-Body Therapies

**Figure 5:**
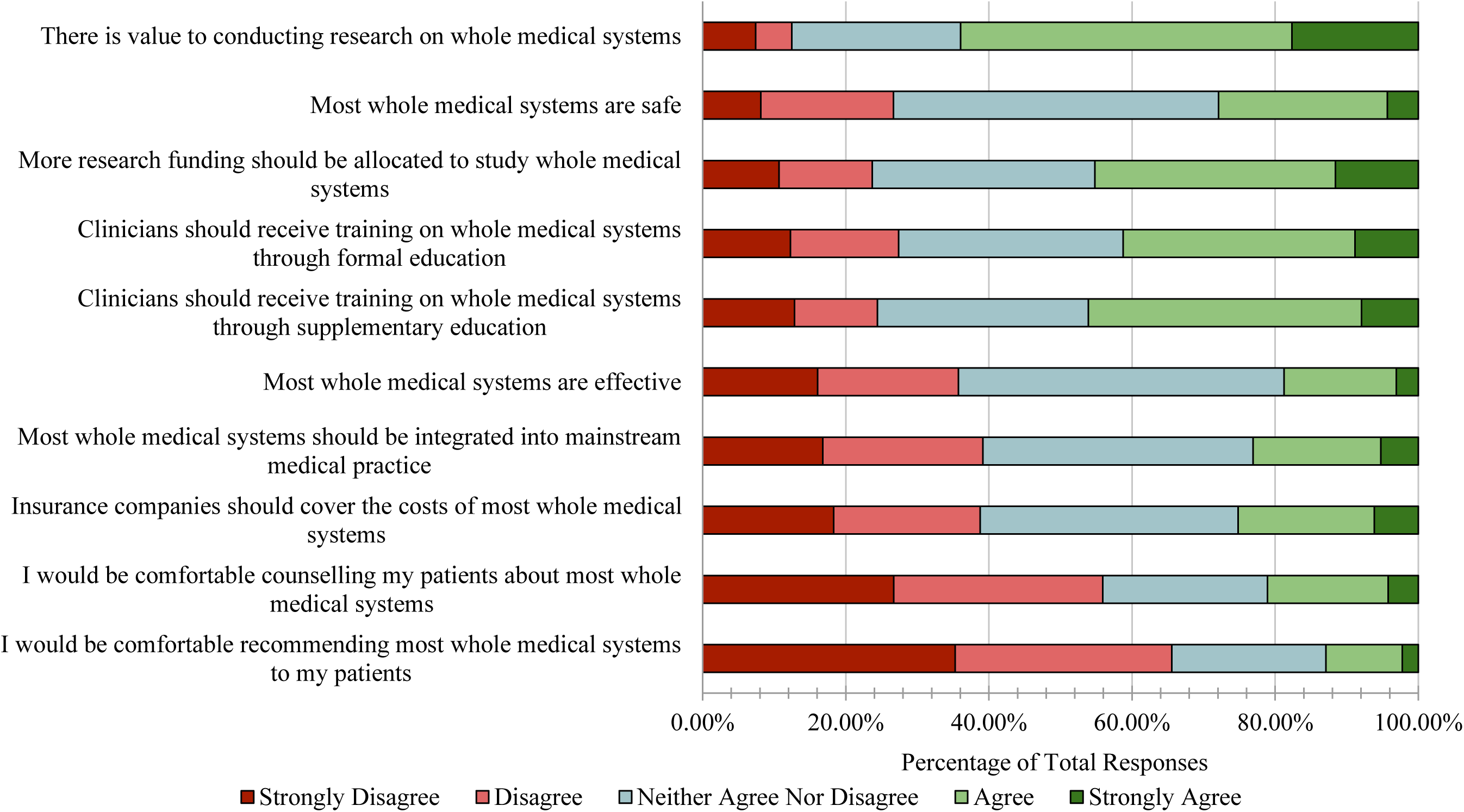
Agreement with the Following Statements Regarding Whole Medical Systems

**Figure 6:**
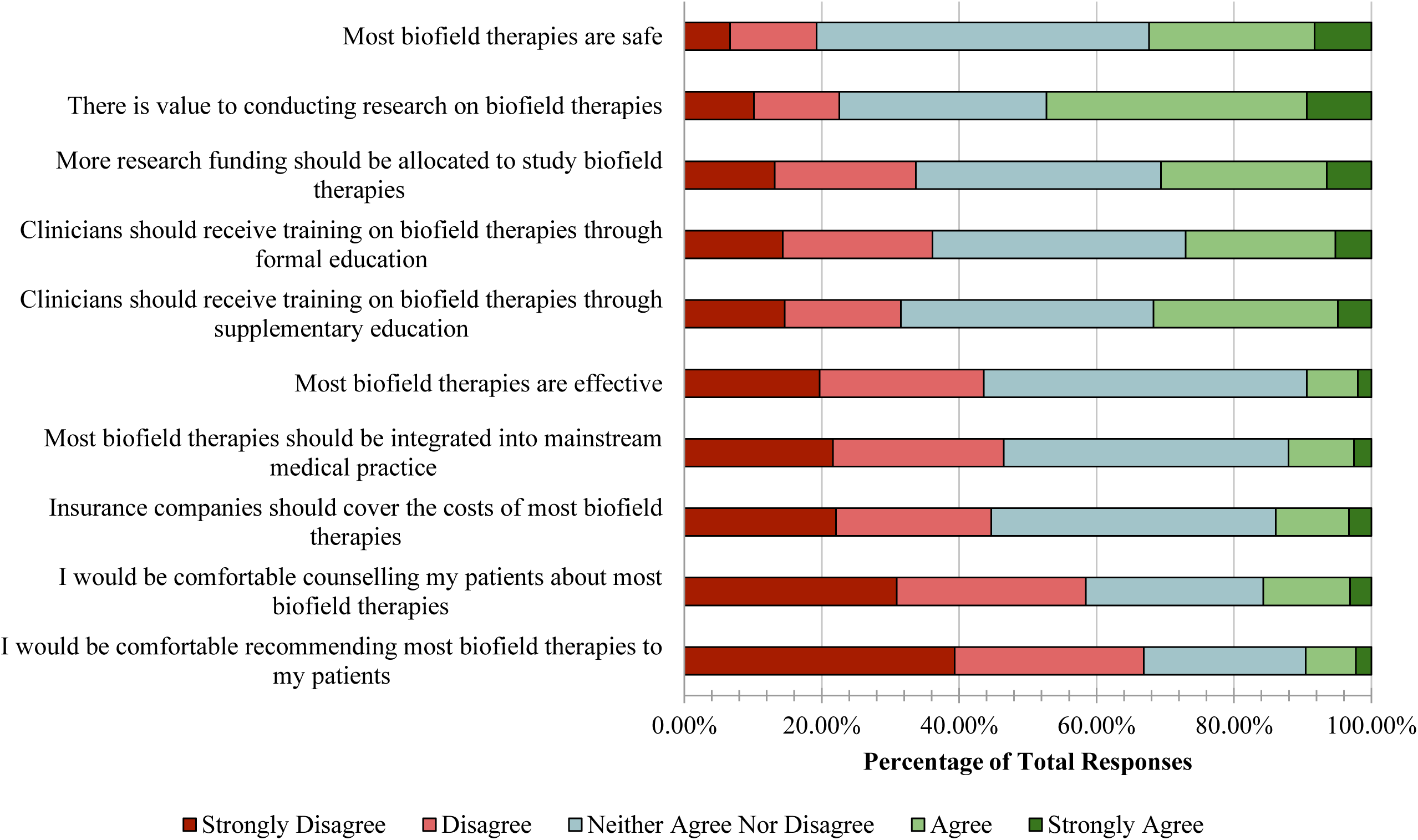
Agreement with the Following Statements Regarding Biofield Therapies

**Figure 7:**
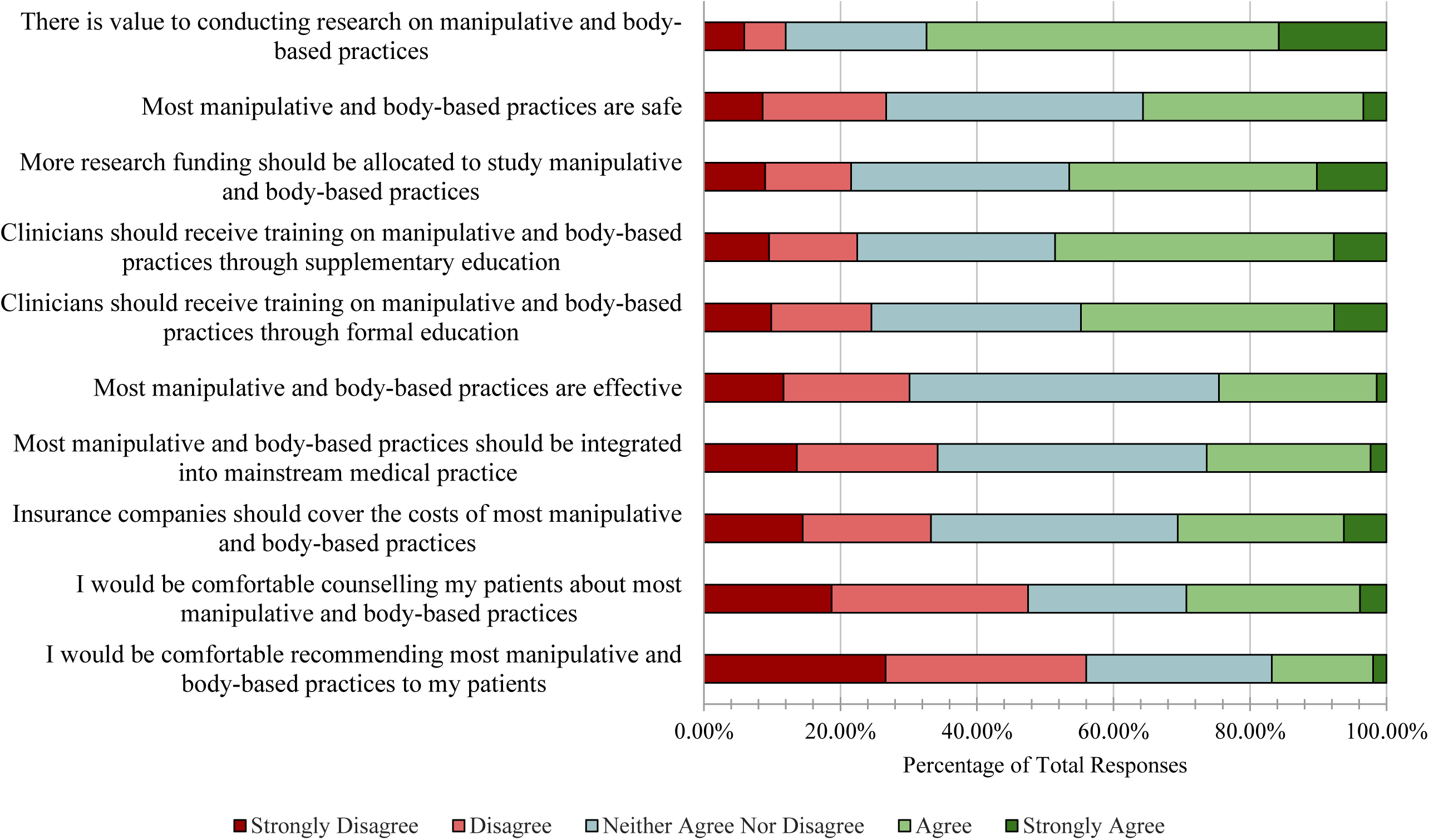
Agreement with the Following Statements Regarding Manipulative and Body-Based Practices

**Figure 8:**
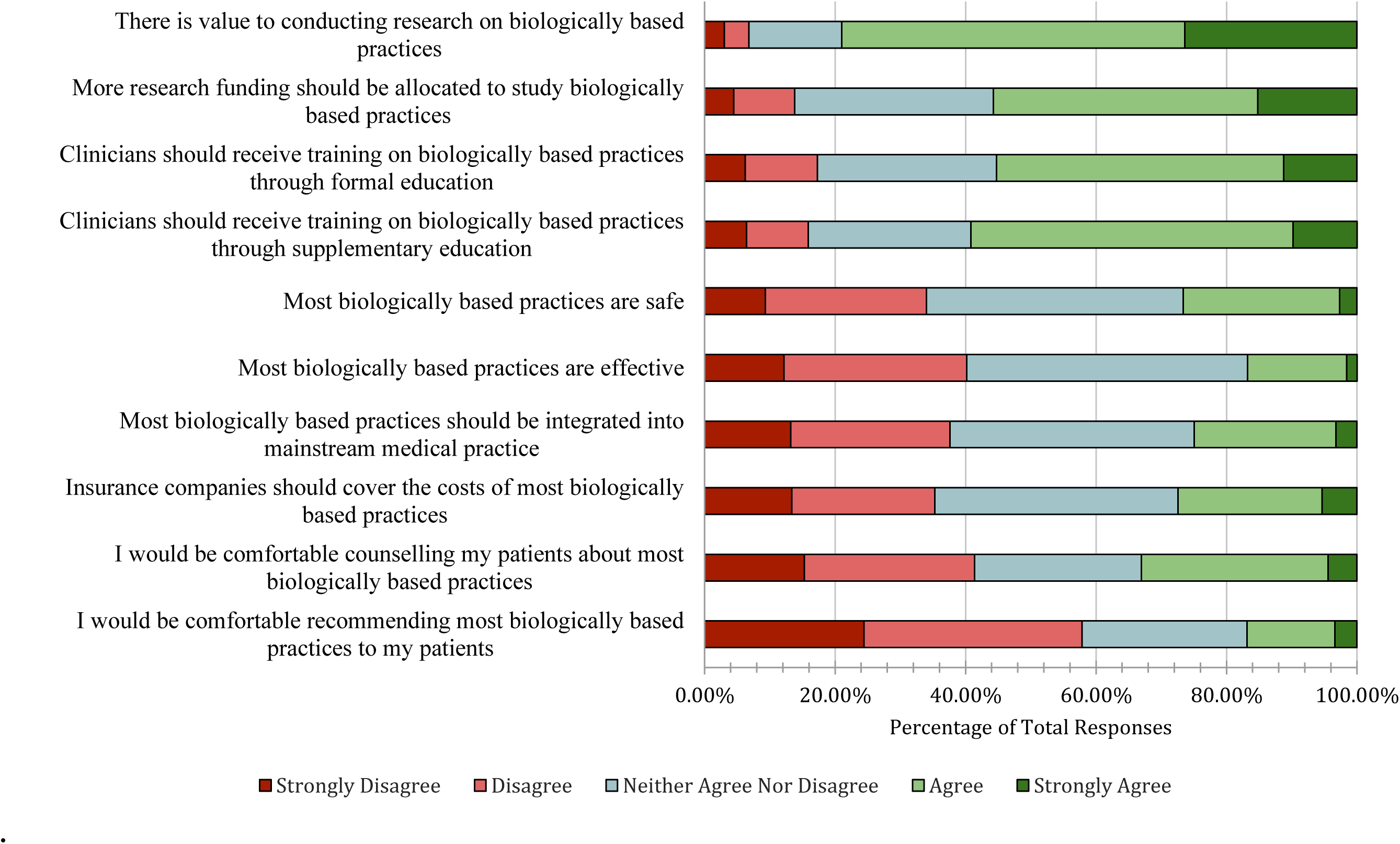
Agreement with the Following Statements Regarding Biologically Based Practices

### Thematic Analysis

A total of 129 responses were grouped into 52 codes that were further grouped into 7 themes. These themes represent distinct patterns found within the data [31]. The most common theme across all responses was the “need for further research on CAIM”. This was presented as a lack of quality and quantity of current CAIM research. Next, “integration and regulation of CAIM within the conventional healthcare system” included the opinions of many respondents who felt CAIM needs more regulation, and that CAIM practices should be integrated into the health system. Next, “patient safety, legal and ethical concerns” reflects respondent concerns about deceiving patients, spreading misinformation, fostering false hope, replacing conventional medical therapies and compromising safety in infants, and believing that those who provide CAIM should face legal consequences. Next, “diverse perspectives of CAIM” encompassed the varying perspectives respondents presented such as supporting or opposing certain CAIM practices. Next, “observational insights and outcomes of CAIM use” included insights and outcomes respondents had observed in a clinical or personal setting. For example, some respondents observed patient improvement following CAIM use, interest in CAIM, and increase in CAIM use following the COVID-19 pandemic. Next, the theme of “education and training for clinicians” was reflected by respondent beliefs that healthcare clinicians and researchers need to be familiar with CAIM practices and support for physicians receiving adequate, appropriate, and evidence-based training on CAIM. Lastly, “cultural and personal considerations’’ were presented as the support or opposition of the integration of religious practices and the consideration of cultural support of CAIM. Please see **Table 4** for themes and examples. Coding and thematic analysis data are available at: https://osf.io/729cv

**Table 4:** Representative Quotes from the Thematic Analysis of CAIM Perceptions.

| **Themes** | **Definition and Example** |
| --- | --- |
| **Need for Further Research on CAIM** | This term refers to the pediatric researchers and clinicians' perceptions that there needs to be further research on complementary, alternative and integrative medicine.<br>**"It should be studied.** It should be covered in medical training but not in a way that implies safety, utility, or benefit unless there is strong reproducible evidence that it works. **We need to know about it because these practices are commonly used and will interact with health care including the care I provide. I need to understand that.** I am not sure if in the end it has any added value or not. I am open to finding out. I would love to have alternatives that contribute to better outcomes. Right now, there is very little data to support that, but a lot of sales based on beliefs that have not been investigated." |
| **Integration and Regulation of CAIM within the Conventional Healthcare System** | This term refers to the pediatric researchers and clinicians' perceptions surrounding the integration and regulation of CAIM within the conventional healthcare system.<br>"I agree with **integrating or complementing CAIM with conventional care**, but I **strongly disagree** with using **CAIM as an alternative** to conventional care. |
| **Patient Safety, Legal, and Ethical Concerns** | This term refers to the pediatric researchers and clinicians' patient safety, legal and ethical concerns surrounding CAIM.<br>"There is a real **potential to exploit patient anxiety and lack of information**, and these alternatives sometimes replace scientifically valid medical therapies, rather than **complementing them.**" |
| **Diverse Perspectives of CAIM** | This term refers to the diverse perspectives of pediatric researchers and clinicians' on CAIM.<br>**"I believe CAIM is important mainly for patients with functional disorders.** When patients have serious medical conditions (for example severe IBD) and they receive CAIM instead of conventional treatment, it may be very harmful. **It may be helpful as supplementary treatment for serious diseases**, but mainly for functional disorders." |
| **Observational Insights and Outcomes of CAIM Use** | This term refers to the pediatric researchers and clinicians' observational insights and outcomes of CAIM use.<br>"CAIM has come up in a research study I did ... **A significant portion of respondents indicated they want more information on CAIM.** Since **there is a patient demand**, I think the scientific and research community should respond - first with good science to see what works |
|  | and what doesn't, and then with strategic Knowledge Translation to ensure the public understands.” |
| **Education and Training for Clinicians** | This term refers to the pediatric researchers and clinicians’ perception on CAIM education and training for clinicians.<br>“I would **love to see CAIM integrated into mainstream health**. The issue I see is that **clinicians go and attend a weekend course on CAIM** i.e. hypnosis/acupuncture and then **feel they are qualified enough** to start practicing. **This dilutes the effectiveness** of the CAIM practice.” |
| **Cultural and Personal Considerations** | This term refers to the pediatric researchers and clinicians’ cultural and personal considerations surrounding CAIM.<br>“**I personally only find the alternative healthcare helpful for chronic conditions for myself**. I use the cranial osteopath for various health complaints, herbal supplements for better sleep, vitamins (D3, and B complex) for preventative health, and I only eat fresh home cooked food. I find that family doctors often think about what medicine they can prescribe but often the problem needs more holistic treatment to solve it rather than add another problem. For example, for reflux management, I don't use PPIs due to their side effects, and I manage it with lifestyle choices...” |

## Discussion

The objective of this study was to investigate the perceptions of published pediatrics authors on CAIM. The results of this large-scale, international cross-sectional survey allow for a comprehensive understanding of pediatrics authors’ perceptions towards the use of CAIM. These findings highlight that pediatrics clinicians and researchers have varying perceptions towards CAIM therapies. There were some CAIM therapies such as mind-body therapies toward which respondents had generally positive perceptions. However, respondents had comparatively fewer positive perceptions towards whole medical systems and biofield therapies as compared to mind-body perceptions.

### Comparative Literature

Our findings align with preexisting literature on healthcare clinicians’ and researchers’ perceptions of CAIM. There are three similar studies which investigate clinician and researcher perceptions of CAIM within the fields of oncology, neurology, and psychiatry [32–34]. Respondents in these fields generally were not comfortable recommending and counselling patients about CAIM but agreed that clinicians should receive more formal and supplemental research on CAIM and that more research funding should be allocated for CAIM. Specifically, mind-body therapies were the most favourably viewed CAIM therapies by clinicians and researchers across these three fields. The respondents were most comfortable recommending and counselling patients on these mind-body therapies. Interestingly, our study found that pediatrics clinicians and researchers were neutral (neither agreed nor disagreed) towards the safety of CAIM practices as opposed to oncology, neurology and psychiatry clinicians and researchers in which respondents were most likely to agree that CAIM therapies were safe [32–34].

Our thematic analysis found a key theme of cultural and personal considerations of CAIM. A study exploring the perspectives of pediatrics healthcare clinicians and researchers (HCCRs) about CAIM found that Indonesian HCCRs were more likely to recommend most CAM therapies as compared to Dutch HCCRs [39]. Our demographic data shows only 14.65% of respondents were from South-East Asia and the Western Pacific regions. These regions may see CAIM therapies in a much more positive perception and this can influence the comfort levels that respondents from these regions have in recommending CAIM therapies [35]. Additionally, individuals from South-East Asia are more likely to seek out CAIM training and this can further enhance their positive perceptions of CAIM [35]. In previous literature assessing attitudes of Swiss, American, and German pediatricians and pediatrics subspecialists such as pediatrics oncologists, a common theme of preferring to receive more CAIM education was found in 82%, 84% and 86% of respondents, respectively [36–38]. These results were seen in pediatrics residents as well, where 88% of respondents reported preferring to increase their knowledge of CAIM [39]. Similarly, our results demonstrate that respondents agree clinicians should receive formal and supplementary education across almost all CAIM therapies. These survey responses align with thematic findings which highlight a theme of a ‘lack of education and training” regarding CAIM. Further research should focus on tailored educational resources specific to pediatrics clinicians and researchers to be reflective of the pediatric patient CAIM interest and use.

Our results highlighted that some perceived challenges with CAIM include a lack of integration within mainstream healthcare systems. Pediatricians have reported that they rarely ask patients about their CAIM use [40]. As CAIM therapies can interact, antagonistically or synergistically, with conventional medications [41,42], physicians must be open to initiating a conversation about CAIM therapies with their patients to ensure they are aware of all the therapies that the patient may be pursuing. A previous study exploring pediatrician experiences with CAIM therapies show that pediatricians most commonly referred patients to mind-body therapies [18]. This could help explain why the most preferred CAIM therapy in our study was mind-body therapies and why our respondents were most comfortable recommending and counselling patients about mind-body therapies such as biofeedback. Pediatricians are significantly less likely to recommend manipulative and body-based practices such as chiropractic therapy as compared to general physicians [43]. This could explain why respondents in our study were unsure about the safety and efficacy of manipulative and body-based practices and why they were hesitant to recommend and counsel patients about these practices.

### Strengths and Limitations

A strength of this study is that by exploring the perceptions of pediatrics researchers and clinicians using a large, international sample of participants with varying opinions on the topic, the findings of this study will be generalizable. Moreover, emails were sent to researchers and clinicians who published an article recently. By doing this, we limited the number of invalid or inactive email addresses. There was also a high survey completion rate among those who initiated the survey. This study also has some limitations. First, our sample may only include individuals who can understand English, as the survey and email were sent in English. Therefore, this study might not capture the perceptions of non-English-speaking researchers and clinicians. Secondly, given the self-reported, cross-sectional nature of the survey, our analysis is subject to recall bias, which refers to the varying accuracy and completeness with which different participants remember information and experiences, in this case regarding CAIM [44]. Thirdly, the categorization of CAIM therapies on the survey was broad, allowing for the grouping of various therapies. While this allowed for a generalized approach, many respondents expressed that the categorization needs to be narrower. For example, manipulative and body-based practices cover massage, chiropractic therapy, reflexology etc. As such, future work should explore perceptions of specific therapies. It is also important to note that perceptions towards CAIM modalities may be due to a variety of concerns ranging from lack of evidence and safety to a lack of knowledge or training and future research should explore concerns surrounding specific perceptions. Unfortunately, as responses were collected anonymously, it is not possible to analyze the characteristics of those who participated versus did not participate in our survey. Additionally, the phrasing "most CAIM therapies" in some survey questions may have introduced a bias toward more negative responses. Instead, if participants were asked about "CAIM therapies with a certain level of evidence", responses might have been more positive. This highlights the influence of wording on survey results and should be considered when interpreting our findings. Finally, the study also contains non-response bias, especially with a low response rate, as authors in our initial sample who declined participation may have answered the questions in our survey differently, resulting in a potential difference between participants and non-participants [44].

## Conclusion

This study aimed to investigate the perceptions of both pediatrics researchers and clinicians regarding CAIM. Our analysis revealed that respondents had varied perceptions of CAIM therapies; mind-body therapies were the most favourable and biofield therapies had the least favourable perception. In general, pediatrics physicians and clinicians lacked comfortability in recommending and counselling CAIM therapies. Out of all the categories of CAIM therapies, respondents were most comfortable with mind-body therapies. There was also a sentiment of not having enough formal or supplemental evidence-based training in CAIM. Previous literature suggests that pediatrics residents, pediatricians and pediatrics subspecialists overwhelmingly prefer to have more CAIM education and knowledge to better serve their pediatric patients. These findings support the development of evidence based CAIM training and educational resources through further research.

## Data Availability

All data and materials associated with this study have been posted on the Open Science Framework.

https://doi.org/10.17605/OSF.IO/Z6U4X

## List of Abbreviations

CAIM: complementary, alternative, and integrative medicine
CHERRIES: Checklist for Reporting Results of Internet E-Surveys
HCCRs: Healthcare clinicians and researchers
MEDLINE: Medical Literature Analysis and Retrieval System Online
NLM: National Library of Medicine
OSF: Open Science Framework
PMIDs: PubMed Identifiers

## Declarations

### Human Ethics and Consent to Participate

This study received approval from the University Tübingen Research Ethics board before commencement (REB Number: 389/2023BO2).

### Consent for Publication

All participants provided their consent to have their data collected and included in the results reported in this study.

### Availability of Data and Materials

All data and materials associated with this study have been posted on the Open Science Framework and can be found here: https://doi.org/10.17605/OSF.IO/Z6U4X

## Acknowledgements

None.

